# Prevalence and associated factors of khat chewing among pregnant women: A Systematic Review and Meta-analysis

**DOI:** 10.1101/2022.04.21.22274111

**Authors:** Muluken Basa, Catherine Comiskey

**Affiliations:** School of Nursing and Midwifery, Trinity College Dublin, Dublin University, Dublin, Ireland; School of Public Health, College of Medicine and Health Sciences, Arba Minch University, Arba Minch, Ethiopia

**Keywords:** Catha edulis, chat, khat, pregnancy: Ethiopia: Yemen, substance use, associated factor

## Abstract

**Background:** Khat (Catha edulis) is a stimulant plant, broadly cultivated and consumed in the Horn of Africa and the Arabian Peninsula. It contains Cathinone, which is an amphetamines-like chemical and causes various adverse outcomes for pregnant women and babies when it is consumed during pregnancy. Decisive estimates of the prevalence of khat chewing and related risk factors which may increase this practice have not been determined.

**Aim:** To determine the pooled prevalence and associated factors of khat chewing among pregnant women in the Horn Africa and the Arabian Peninsula countries with a view to informing targeted interventions for the region.

**Method:** The study protocol was prepared and registered on PROSPERO, ID CRD42021190837. A database search including Gray literature and Google scholar was explored to identify 667 studies. Finally, 14 studies were considered relevant for meta-analysis, after removing 259 duplicates, 388 unrelated topics and 6 studies with full text examination. The Newcastle-Ottawa Scale quality assessment tool was used to assess the quality of the studies. The pooled prevalence was determined by using the random-effect model and the p- values of ≤ 0.05 were considered stastically significant to examine associations. Statistical heterogeneity amongst the studies was assessed by Cochrane chi-square and the I^2^ statistical test.

**Main Findings:** From the meta-analysis of 14 studies with 15,343 study participants, the pooled prevalence of khat chewing among pregnant women was 21.42%, 95% CI (14.49 - 29.29); (I ^2^=99.05% (p<0.0001). The results of the meta-analysis demonstrated that pregnant women who had a khat chewing partner [OR 6.50 (95% CI 5.01, 8.43)]; low educational status [OR 2.53 (95% CI 2.24 - 2.85)], lived in rural area [OR 1.69 (95% CI 1.52 – 1.88)] or had a low level of income [OR 1.70 (95% CI 1.55 – 1.87)] were significantly more likely to chew khat during pregnancy.

**Conclusion:** The prevalence of khat chewing amongst pregnant women in the Horn of Africa and the Arabian Peninsula has never been measured before and was found to be high. Partners khat chewing status, maternal low educational and economic status were the main factors associated with the problem. Designing intervention strategies to specifically target these risk factors and reduce the burden of the problem for women and their babies is urgently needed.

## Introduction

Khat (Cath edulis) is an evergreen plant widely cultivated and consumed in the eastern Africa and the Arabian Peninsula countries (1, 2). It was originated from Ethiopia, but later distributed to different countries such as in Somalia, Kenya, Malawi, Uganda, Zimbabwe, Afghanistan, Tanzania, the Congo, Zambia, Yemen and Madagascar(3). It also known as *qaat* and *jaad* in Somalia, maria in Tanzania and Kenya and *echat* in Ethiopia (4). It contains an amphetamines-like chemical called Cathinone (5). It is a central nervous system stimulant and gives the chewer a mild high to euphoria (1, 6). Traditionally, the majority of people chewing khat were adult men, khat was a social drug and people gathered together to consume for a social event (5). However, this trend has been changing, currently, people from different age groups including women consume this plant for recreational and other purposes (7).

Individual regional study findings indicate that the magnitude of khat chewing in the Horn of Africa and the Arabian Peninsula countries were distributed unevenly (2, 8). The higher prevalence was observed in Yemen, with 90% adult males and around 73% of women chew khat (8). Almost similar proportion, of 90% and 88%, khat chewing prevalence were reported in Djibouti and northwestern Kenya, respectively (8). In contrast, population-based survey conducted in southwest Ethiopia and Uganda identified a prevalence of 32%-38.6 %, in the general population (9, 10). A similar study conducted in the Jazan Region, Saudi Arabia, identified that the lifetime prevalence of khat chewing was 33.2% in the general population (3).

Moreover, the national population survey in Yemen stated that 40.7% of women chewed khat during their last pregnancy (11). Similarly, a community-based study conducted in rural parts of eastern Ethiopia identified a prevalence of 34.6 % among pregnant women (12). In comparison, the prevalence was found to be 19.5% in southwest Ethiopia in the same population group (13). The proportion of the problem was known to vary across the regions, with the lowest prevalence of 10% and the highest prevalence of 37% registered in the southern and eastern region of Ethiopia, respectively (14, 15).

Individual studies in a range of regions have identified that factors contributing to khat chewing during pregnancy include having a lower level of education and socioeconomic status, living in mountainous and rural areas, ethnicity and religious belief, number of children, having a partner who chews khat, physical and mental health problems (2, 11, 13, 16-18). Nakajima (2017) identified recreation and socialization as the main motivational factors of khat chewing during pregnancy. Alcohol drinking, cigarette smoking, and the use other substances were also the most important predictors of khat chewing (13, 18-21).

Khat chewing during pregnancy has many adverse outcomes both for the mother and the fetus/ baby (6, 22, 23). Khat chewing mothers are at high risk for blood transfusion, breech presentation, anemia, Premature Rupture of Membrane (PROM), Post Partum Hemorrhage (PPH), perineal tears, Intra Uterine Fetal Death (IUFD), depression, loss of appetite, anemia, sleep disturbance and memory impairment (2, 5, 13, 15, 24-27) (28).

Even though the problem is highly prevalent in the Horn African countries and the Arabian Peninsula, little has been reported about the overall prevalence and associated factors among pregnant women. The aim of this systematic review to answer the following study questions; What is the pooled prevalence of khat chewing among pregnant mothers in the Horn Africa and the Arabian Peninsula and what are the main contributing factors associated with khat chewing during pregnancy? The results of this study will provide specific guidance and direction for policymakers and practitioners in the design of targeted interventions to address this challenge. Furthermore, it also provides input and direction for future research in the area.

## Methods

### Search and selection strategy

A systematic search technique was applied to the electronic databases MEDLINE, CINAHL, EMBASE, and PubMed without a year of publication and language restriction on June, 2021. The keyword identified for the main term khat included “Khat” OR “Cathas” OR “Catha edulis” OR “Mira!”, and for pregnant mother “Pregnant Wom!n” OR “perinatal” OR “Expectant mother” OR “Expectant woman” OR “Prenatal” OR “pregnancy”.

In addition, grey literature searches and manual google scholar searches were also applied. Finally, the search results were collected in Endnotex9 and then exported into Covidence(29). Two independent authors selected literature based on selection criteria using Covidence (29). The Preferred Reporting Items for Systematic reviews and Meta-Analyses (PRISMA) was used to describe the process of study selection (30). see Figure 1.

**Figure 1:**
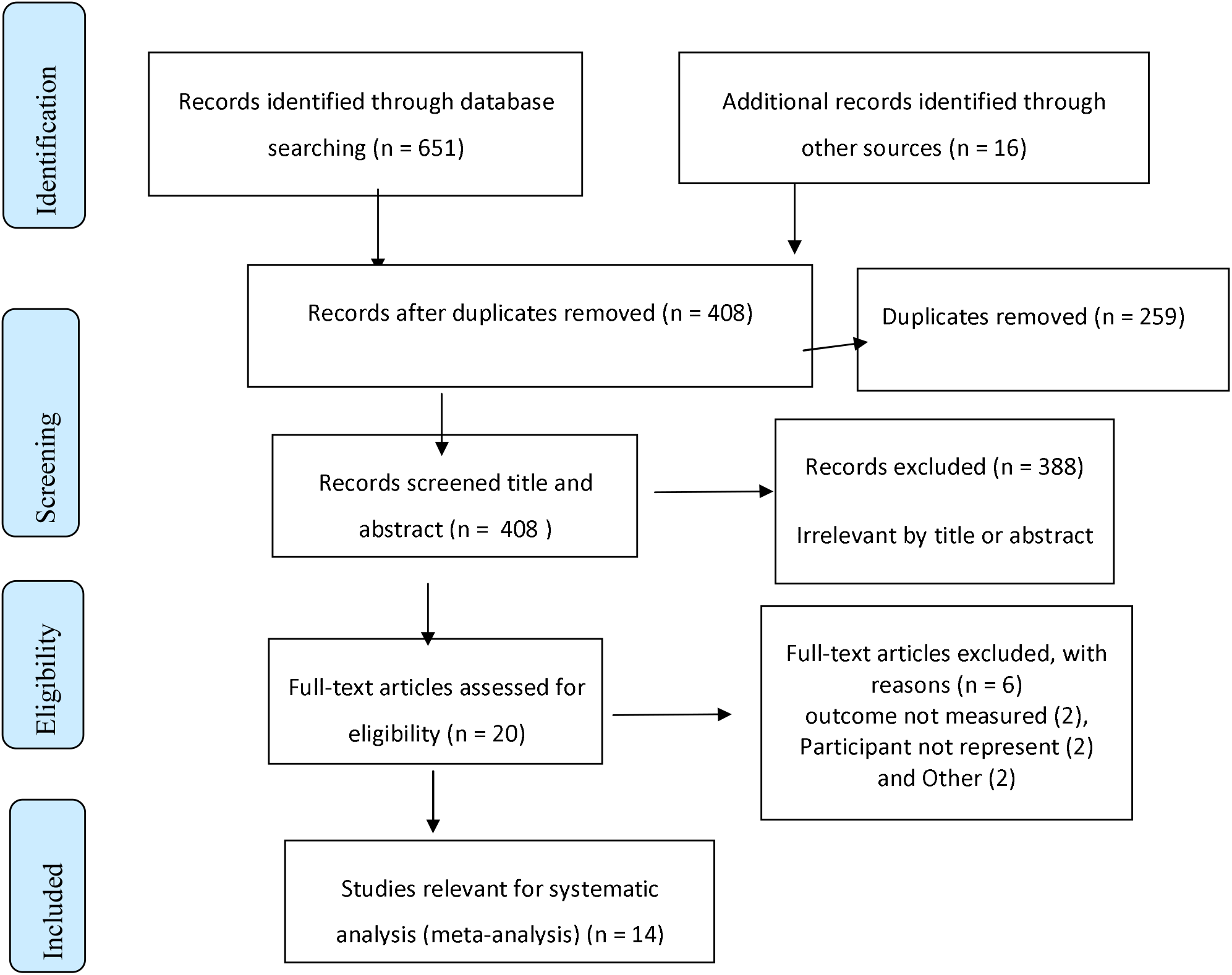
PRISMA flowchart describing the selection of studies for the systematic review and meta-analysis of prevalence and associated factors of khat chewing among pregnant women in Horn Africa and the Arabian Peninsula, 2021.

### Inclusion/Exclusion criteria

#### Inclusion criteria

✓ **Population;** studies conducted on pregnant women/ during a pregnancy.
✓ **Intervention;** studies included khat chewing habits.
✓ **Outcome**; the outcome of the studies included was the proportion of khat chewing and associated factors: socio-demographic, emotional, mental, and physical health-related factors.
✓ **Study design**; all quantitative studies that contain the number/ proportion of khat chewing during pregnancy and/or associated factors were included in the study.
✓ **Study area;** studies conducted in in the East Africa and the Arabian Peninsula countries were included in the study.

#### Exclusion criteria

✓ Literature with undefined/ unrelated outcome measures, articles with missing or insufficient outcomes were excluded from the systematic review. Literature reviews, individual case reports and patient stories were also excluded.

### Outcome measures

The two main outcomes of this study were 1) to the determine the pooled prevalence of khat chewing among pregnant women in the Horn Africa and the Arabian Peninsula countries and 2) to identify the associated factors of khat chewing during pregnancy.

Khat chewing during pregnancy, in this study was defined as the chewing of any amount of khat at least once during the current pregnancy or during a previous pregnancy if the women was not currently pregnant.

### Quality assessment

The Newcastle-Ottawa Scale (NOS) quality assessment tool was used to assess the quality of the studies (31). The tool has 8 quality indicators which divided in 3 sections. Then, studies were classified as high (7-8), medium (4-6) and low (1-3) quality, based on their respective quality score (32). see supporting document 1. The investigators evaluated the quality of the papers independently and then met to discuss and agree on individual scoring differences. Finally, eight studies were scored a medium quality and six scored high-quality study. See table 1.

**Table 1:** Summary characteristics of included studies for the systematic review and meta-analysis of prevalence and associated factors of khat chewing among pregnant women in Horn Africa and the Arabian Peninsula, 2021.

| s. no | Author/Year | Study location | Study design/ setting | weight |  | Participants | Prevalence (95%CI) | Respo. rate | Quality |
| --- | --- | --- | --- | --- | --- | --- | --- | --- | --- |
|  |  |  |  | Fixed | Random |  |  |  |  |
| 1 | (Mekonnen et al. 2020) | Bale zone, Oromia region, Ethiopia | case-control / Institution | 2.67 | 7.11 | 409 | 12.7% (9.84-16.37) | 96.7% | MEDIUM 6 |
| 2 | (Woldetsadik et al. 2019) | Bale Zone, Oromia region, Ethiopia | cross-sectional / community | 4.84 | 7.17 | 743 | 10.4% (8.32-12.7) | 99.3% | MEDIUM 6 |
| 3 | (Tesfay et al. 2019) | Jimma town, Oromia region, Ethiopia | case-control / Institution | 2.19 | 7.08 | 336 | 35.7% (30.7-40.95) | 100% | MEDIUM 6 |
| 4 | (Tesso et al. 2017) | Jimma zone, Oromia region, Ethiopia | cross sectional / Institution | 1.91 | 7.05 | 293 | 24.9% (20.2-30.1) | 98.9% | HIGH 7 |
| 5 | (Nakajima et al. 2017) | Jimma zone, Oromia region, Ethiopia | Cress-sectional / Institution | 4.19 | 7.16 | 642 | 19.15% (16.25-22.34) | 97.3 | HIGH 7 |
| 6 | (Demelash et al. 2015) | Bale zone, Oromia region, Ethiopia | case-control / Institution | 2.53 | 7.1 | 387 | 12.4% (9.43-15.97) | 94 % | MEDIUM 6 |
| 7 | (Mekuriaw et al. 2020) | Gedeo zone, Sothern NNPRS, Ethiopia | Cross-sectional / Institution | 4.68 | 7.17 | 718 | 9.9% (7.86-12.24) | 97.9 | HIGH 7 |
| 8 | (Alamneh et al. 2020) | Gurage Zone, Sothern NNPRS, Ethiopia | Cross-sectional / community | 2.23 | 7.08 | 341 | 35.8% (30.8- 41.0) | 96.9% | MEDIUM 6 |
| 9 | (Mekuriaw et al. 2019) | Gedeo zone, Southern Ethiopia. | Cross-sectional / Institution | 4.68 | 7.17 | 718 | 10% (7.86-12.24) | 94.6% | HIGH 7 |
| 10 | (Fetene et al. 2021) | (Jigjiga, Dire Dawa and Harar towns). Eastern Ethiopia | Cross-sectional / Institution | 3.33 | 7.14 | 510 | 19.6% (16.33-23.23) | 96.9% | HIGH 7 |
| 11 | (Yadeta et al. 2020) | Harar town, Eastern Ethiopia | cross-sectional / Institution | 11 | 7.22 | 1688 | 37.2% (34.9-39.53) | NA | MEDIUM 5 |
| 12 | (Tesfaye et al. 2020) | Addis Ababa, Ethiopia | cross sectional / Institution | 3.82 | 7.15 | 585 | 10.4% (8.14-13.1) | 98.6% | HIGH 7 |
| 13 | (Khawaja et al. 2008) | 11435 households In Yemen | cross-sectional / Community | 47.82 | 7.25 | 7343 | 40.5% (39.44-41.7) | NA | MEDIUM 6 |
| 14 | (Muhammed 2015) | in Sana'a City Yemen. | Cross-sectional / Institution | 4.11 | 7.16 | 630 | 35.9% (32.2-39.7) | NA | MEDIUM 4 |
|  | Total (fixed effects) |  |  | 100.00 | 100.00 | 15343 | 29.92% (29.20 - 30.66) |  |  |
|  | Total (random effects) |  |  | 100.00 | 100.00 | 15343 | 21.42% (14.49 - 29.29) |  |  |

### Data extraction procedure

A standardised data extraction tool was prepared in Microsoft Excel and general information of the study, participant eligibility criteria, study design, sample size, study results, and other important notes were extracted to it. The most common factors were isolated and summarised in the form of two by two tables for meta-analysis.

### Data Analysis

The summary table was prepared to describe the findings in each study. The pooled prevalence of khat chewing in pregnant women was calculated using MedCalc software-version 20.0.13 and the pooled odd ratios for predictors was calculated using RevMan software version 5.3. The p-value ≤ 0.05 at 95% confidence interval was considered statistically significant. The random-effect model was used to determine the pooled prevalence. Clinical heterogeneity was controlled across the studies with proper selection criteria and statistical heterogeneity of the studies was assessed using the chi-square and the I^2^ statistics test with p-value. I^2^ values of 25%, 50%, and 75% were used to define as low, medium, and high heterogeneity, respectively (33). The Egger’s test was used to assess the presence of publication bias.

## Results

### Search and selection results

The database and manual search strategy was applied and 679 citations with 259 duplicates were found, with the latter removed using Covidence(29). The titles and abstracts of the remaining 408 unique papers were screened, of these, 388 were considered irrelevant as they did not meet the inclusion criteria for the review. Full-texts of the remaining 20 citations were reviewed and six studies which did not contain our study outcome were excluded. Finally, 14 studies with 15343 study participants, were used for systematic analysis (meta-analysis). The search results and details of the selection process was described with a PRISMA diagram in Figure 1(30).

Among 14 studies considered relevant for systematic review meta-analysis, 12 studies were conducted in Ethiopia (six in Oromia region (13, 17, 18, 22, 23, 34), three in Southern region (14, 35, 36), two in Eastern part Ethiopia (15, 16) and one in Adis Ababa (37)) and two studies were conducted in Yemen (11, 27). See table 1.

### Meta-analysis result

#### The prevalence of khat chewing among pregnant women

In this study, a total of 15343 study participants were included from 14 studies. This study confirmed the pooled prevalence of khat chewing during pregnancy, using random-effect model, was 21.42% with a 95% CI (14.49 - 29.29). See table 1. The result detected statistically significant heterogeneity among the studies (I^2^ =99.05% (p<0.0001). The Egger’s test indicates that publication bias was not statistically significant (p =0.0034) See figure 2.

**Figure 2.**
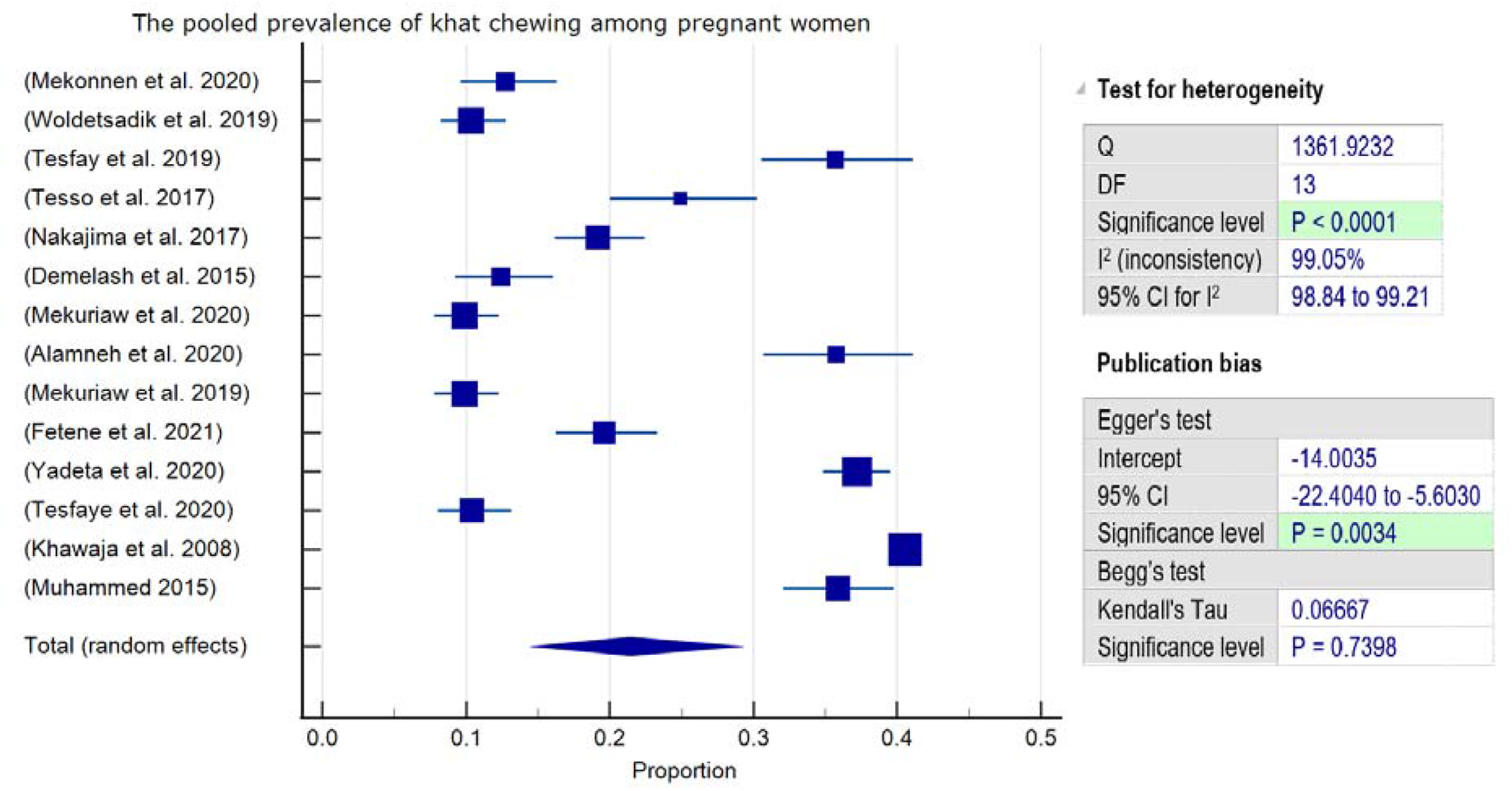
Forest plot of the pooled prevalence of khat chewing among pregnant women in Horn Africa and the Arabian Peninsula, 2021.

#### Sub-group analysis

The sub-group analysis was done to determine pooled prevalence of khat chewing in Ethiopia. Among 12 studies with 7270 participants the prevalence was found to be 18.91%, with the 95% CI (12.75 to 25.96). The analysis demonstrated significant heterogeneity between the studies (I ^2^ =98.12%: p<0.0001). The result of Egger’s test did not demonstrate a statistically significant publication bias (p =0.356). see Figure 3.

**Figure 3.**
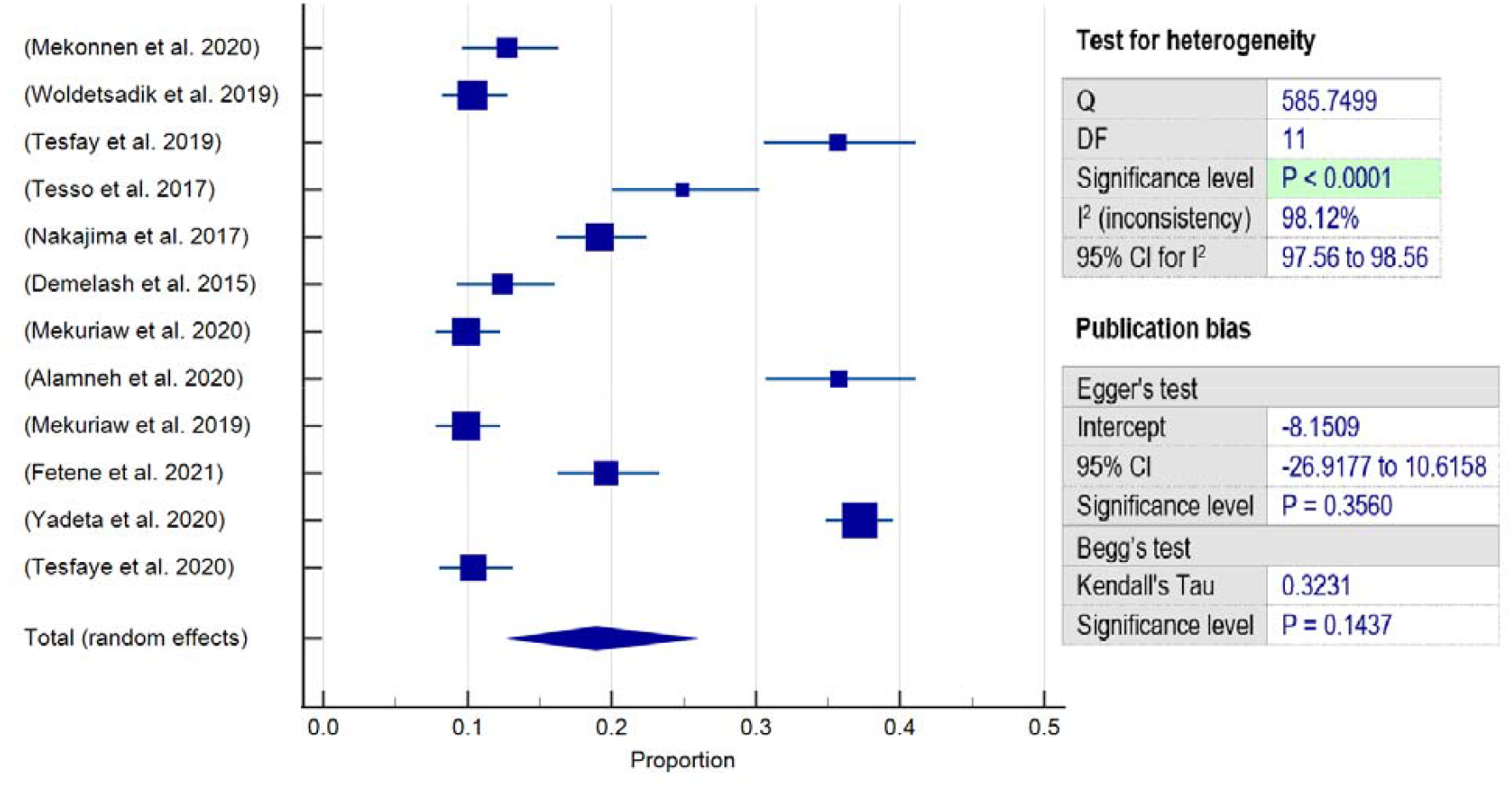
Forest plot of the sub-group analysis on pooled prevalence of khat chewing among pregnant women in Ethiopia, 2021.

**Figure 4:**
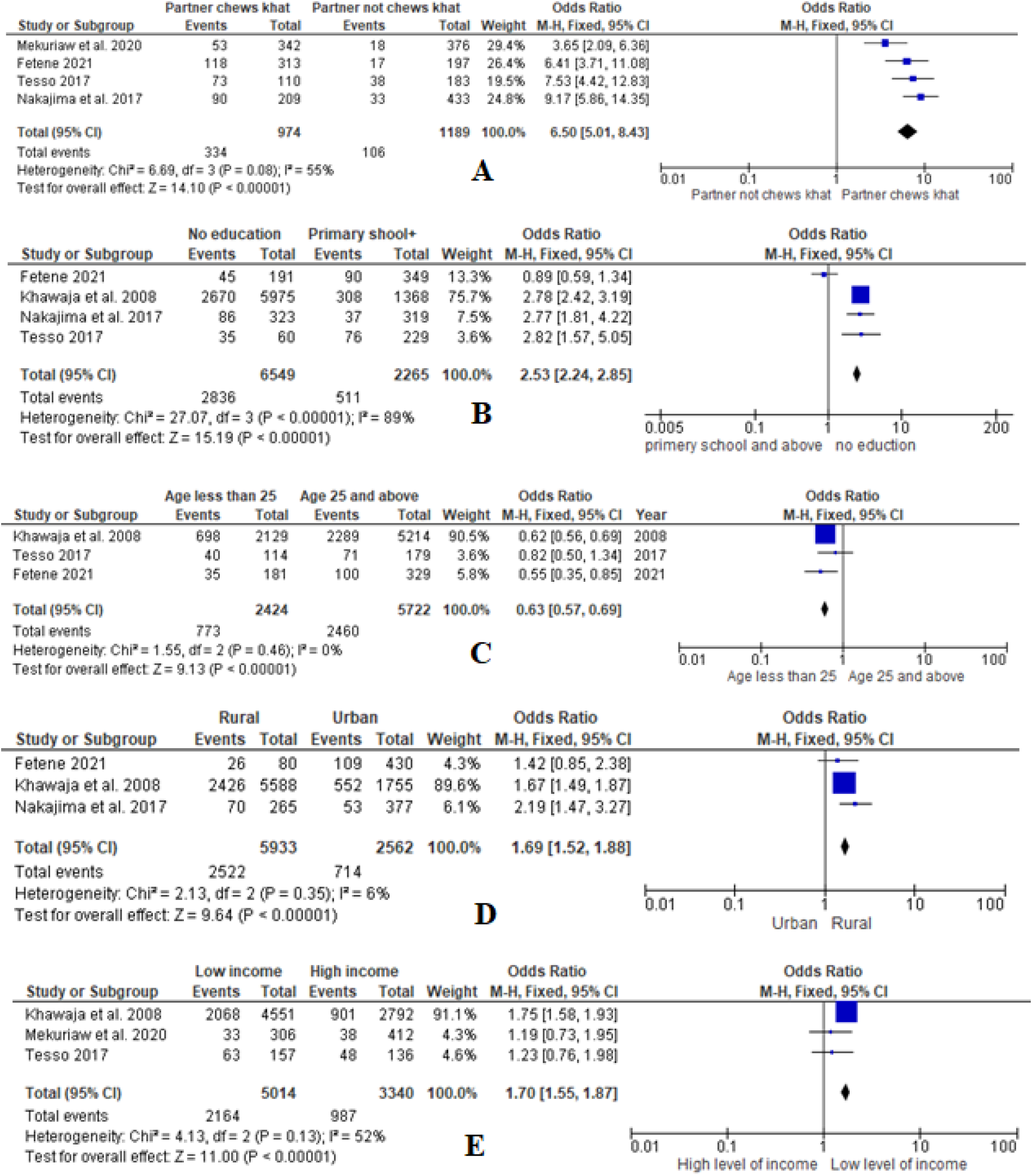
Forest plot of factor association of khat chewing with (A) Partener khat chwing status B) maternal education C) Age D) Place of living E) Level of income) among pregnant women, 2021.

Among sub group-analysis in Ethiopian regions, the highest prevalence of 28.09%, with the 95% CI (12.81 - 46.59), was registered in the Eastern part of Ethiopia and the lowest prevalence of 17.13%, 95% CI (5.99 - 32.44), was observed in southern region of Ethiopia. The pooled prevalence of khat chewing in Oromia region of Ethiopia was 18.51%, 95% CI (12.05 - 25.99). see table 2.The overall prevalence of khat chewing among pregnant mothers in Yemen found to be 38.58% with the 95% CI (34.11 - 43.14). see table 2.

**Table 2.** Table of the sub-group analysis on pooled prevalence of khat chewing among pregnant women in Ethiopian regions and Yemen, 2021

| Region | Number of studies | Participant | Prevalence (95% CI) | Heterogeneity |
| --- | --- | --- | --- | --- |
| Oromia, Ethiopia | 6 (13, 17, 18, 22, 23, 34) | 2810 | 18.51% (12.05 - 25.99) | P < 0.0001, I<sup>2</sup> 95.69% |
| Southern Ethiopia | 3 (14, 35, 36) | 1777 | 17.13% (5.99 - 32.44) | P < 0.0001, I<sup>2</sup> 98.24% |
| Eastern Ethiopia | 2 (15, 16) | 2198 | 28.09% (12.81 - 46.59) | P < 0.0001, I<sup>2</sup> 98.35% |
| Overall Yemen | 2 (11, 27) | 7973 | 38.58% (34.11 - 43.14) | P = 0.0207, I<sup>2</sup> 81.32% |

### Factors associated with khat chewing during pregnancy

This meta-analysis identified significant factors contributing to khat chewing during pregnancy. In summary these included maternal educational status, place of residence, level of income, partner khat chewing status, and maternal Antenatal Care (ANC) follow-up. Whereas, lifetime prevalence of maternal use of alcohol did not demonstrate a statistically significant association. Figure 3:A-E

Partener khat chwing status found to the major pridictor. Those mothers who had a partner who chewed khat were more than 6.5 times more likely to chew khat than there counterparts [OR 6.50 (95% CI 5.01, 8.43); I^2^ =55%; p<0.08]. Heterogeneity among the studies was medium and not stastically significant (figure 3A).

Pregnant mothers who did not have formal education were two and a half times more likely to chew khat during pregnancy than mothers who attained at least primary school (95% CI 2.24 - 2.85) see Figure 3B. High level of statistical heterogeneity was observed between studies (I^2^=89%; p<0.00001).

Maternal place of living was also found to be a significant determinant of khat chewing, mothers living in rural regions were 1.7 times more likely to chew khat during pregnancy than mothers who lived in urban areas (95% CI 1.52 – 1.88)(I^2^ =6%; p<0.36) figure 3D. similarly, those pregnant mothers who had an income status of low were 1.7 times more likely to chew khat than high-income mothers (95% CI 1.55 – 1.87). See Figure 3E.

Mothers who did not have ANC follow-up either in current or during last pregnancy were more than one and a half times more likely to chew khat than their counterparts (OR = 1.53; 95% CI 1.39–1.68;) (see Figure 3F). Maternal ever alcohol use did not show a statistically significant association in pooled odd ratio.

## Discussion

The aim of this study was to determine for the first time, the pooled prevalence and associated factors of khat chewing among pregnant women in Horn Africa and the Arabian Peninsula countries with a view to informing targeted interventions for policy and practice. It is recognised that substance use during pregnancy can result in multiple physical, emotional, social, and psychological problems (2, 38, 39). However, it is also recognised that women chew the substance khat during pregnancy for various reasons including relief from stress, enjoyment with their partners, to feel happy, to socialise and for habitual and medicinal reasons (13, 14).

The pooled prevalence of khat chewing amongst pregnant women in the Horn of Africa and the Arabian Peninsula was found to be 21.42%, with 95% CI (14.49 - 29.29). The prevalence of khat chewing was known to be variable across the countries, regions and population groups (21, 40). Similarly, our sub-group analysis showed, the pooled prevalence of khat chewing in Yemen was highest at, 38.58% with the 95% CI (34.11 - 43.14), and this was found to be approximately twice the prevalence in Ethiopia at, 18.91% with the 95% CI (12.75 to 25.96). Among Ethiopian regions: the highest prevalence was observed in the Eastern part at 28.09% with the 95% CI (12.81 - 46.59), and almost a similar proportion of 18.51% with the 95% CI (12.05 - 25.99) and 17.13% with the 95% CI (5.99 - 32.44), was observed in Oromia region and Southern region, respectively. These variations across the countries and regions may be as a result of the difference in population characteristics including religion, geographic area and the distribution of associated factors among the groups (7, 40). For example, the higher prevalence observed in Yemen and in Eastern parts of Ethiopia was may be due to cultural and social reasons as within these regions it is known that the majority of people were of the same religion (21, 40). Different studies have identified that religion is a significant correlate of khat chewing (11, 13).

If we compare our results with studies exploring other socially acceptable drugs in the broader region we see that, our findings are similar to the prevalence of alcohol use among pregnant women in Nigeria, South Africa, Australia, New Zealand and the UK (41-43). The overall prevalence of alcohol drinking among women in Ethiopia was 35.3 %, with less than 1% in the Somalia region and 71 % in the Tigray region (EDHS 2016). Clearly the need to address the use of socially accepted substance use during pregnancy is important for policy and planning in the region.

This systematic review and meta-analysis confirmed age, educational status, place of residence, income, partner khat use, and ANC follow up history were statistically significant predictors of khat use during pregnancy. The systematic review and meta-analysis performed among university students and a cross-sectional study conducted based on Ethiopian Demographic and Health Survey (EDHS) 2016, also found similar variables as predictors of khat use within those populations (21, 40, 44). However, alcohol use, which had been frequently cited as a correlate of khat use, did not show statically significant association in our study (35, 40, 45). This difference may be due to the variation in the study population and setting or for other reasons and further research on this topic among women who are pregnant is warranted.

Pregnant women who had khat chewing partners were over six times more likely to chew khat than those women whose partners did not chew. This result is also supported by the cross-sectional study which assessed the most common reason of khat chewing during pregnancy and the systematic review and meta-analysis conducted among university students (14, 45).

Disadvantaged women in terms of educational status, place of residence and income level were more likely to chew khat than their counter group. The EDHS 2016 results and the World Health Organisation stated that illiteracy and low educational status are one of the main predictors of substance use during pregnancy (44, 46). This may be due to the fact that mothers who have low educational status and live in rural area may have poor knowledge about the teratogenic effects of substance use during pregnancy (16). Moreover, various studies conducted in Yemen and Ethiopia reported that the majority of women who chew khat were farmers and live in remote areas, where accessibility for media and health information was very poor (11, 13, 40). Furthermore, khat chewing is socially acceptable practice and its deeply rooted with the culture and religion of rural communities (2, 7).

## Conclusion

This study confirmed that the prevalence of khat chewing amongst pregnant mothers was high, nearly one third of pregnant mothers’ chew khat in the study areas.

This meta-analysis identified significant factors contributing to khat chewing during pregnancy. This included maternal educational status, place of residence, level of income, partner khat chewing status, and maternal Antenatal Care (ANC) follow-up. Whereas, maternal ever drink alcohol did not show a statistically significant association.

Despite the fact that khat chewing is known to have harmful effects both for the mother and the baby, little effort has been made so far to measure or address the magnitude of the problem. This may be due to the fact that the problem commonly occurs in rural, remote and disadvantaged societies. Therefore, conducting further community based research that can encompass all contributing factors is important. More importantly, designing evidence based and specifically targeted intervention strategies is needed to decrease the prevalence and impact of khat chewing during pregnancy on both mothers and their babies.

## Data Availability

All data produced in the present study are available upon reasonable request to the authors

## Limitation of the study

The main limitation of this study was the absence of sufficient research in the area. This study was planned to include all studies available in east African and Arabian Peninsula countries. However, the studies that were found during the search strategy were only limited to Ethiopia and Yemen. Moreover, some studies did not investigate all associated factors of khat chewing among pregnant mothers.

## Key points

The prevalence of khat chewing amongst pregnant mothers was high in Ethiopia and Yemen.

The prevalence known to be variable across the regions and population groups.

Pregnant women who had khat chewing partner and disadvantaged women in terms of educational and economic status were more likely to chew khat than the opposite group.

Studies conducted in the area are a few, conducting advanced research in the future is recommended.

The need to address the use of socially accepted substance use during pregnancy is important for policy and planning in the region.

## Declarations

### Ethics approval and consent to participate

Not applicable

### Consent for publication

Not applicable

### Availability of data and materials

Not applicable

### Authors’ contributions

MB developed the study protocol, search strategy and data extraction with the supervision of CC. Both authors (MB and CC) involved in study selection, quality appraisal and data analyses. MB prepared the final manuscript. CC participated in revision, discussion and consensus and approved the final manuscript.

## Acknowledgements

Not applicable

## Bibliography

1. Ageely HMA. Health and socio-economic hazards associated with khat consumption. J Family Community Med. 2008;15(1):3–11.

2. Organization WH, editor Assessment of khat (Catha edulis Forsk). Proceedings of the 34th meeting, Expert Committee on drug dependence; 2006.

3. Mahfouz MS, Rahim B-eEA, Solan YMH, Makeen AM, Alsanosy RM. Khat Chewing Habits in the Population of the Jazan Region, Saudi Arabia: Prevalence and Associated Factors. PloS one. 2015;10(8):e0134545–e.

4. Beckerleg S. ‘Idle and disorderly’ khat users in Western Uganda. Drugs: Education, Prevention and Policy. 2010;17(4):303–14.

5. Alemayehu G, Tewodros G. The Chemistry of Khat and Adverse Effect of Khat Chewing. American Scientific Research Journal for Engineering, Technology, and Sciences (ASRJETS). 2014.

6. Magdum SS. An Overview of Khat. Addictive Disorders & Their Treatment. 2011;10(2):72–83.

7. Mihretu A, Teferra S, Fekadu A. What constitutes problematic khat use? An exploratory mixed methods study in Ethiopia. Subst Abuse Treat Prev Policy. 2017;12(1):17.

8. Patel NB. Khat (Catha edulis Forsk) – And now there are three. Brain Research Bulletin. 2019;145:92–6.

9. Alemseged F, Haileamlak A, Tegegn A, Tessema F, Woldemichael K, Asefa M, et al. Risk factors for chronic non-communicable diseases at gilgel gibe field research center, southwest ethiopia: population based study. Ethiop J Health Sci. 2012;22(S):19–28.

10. Belew M, Kebede D, Kassaye M, Enquoselassie F. The magnitude of khat use and its association with health, nutrition and socio-economic status. Ethiopian Medical Journal. 2000;38(1):11–26.

11. Khawaja M, Al-Nsour M, Saad G. Khat (Catha edulis) chewing during pregnancy in Yemen: findings from a national population survey. Maternal And Child Health Journal. 2008;12(3):308–12.

12. Kedir H, Berhane Y, Worku A. Khat chewing and restrictive dietary behaviors are associated with anemia among pregnant women in high prevalence rural communities in eastern Ethiopia. PLoS One. 2013;8(11):e78601.

13. Nakajima M, Jebena MG, Taha M, Tesfaye M, Gudina E, Lemieux A, et al. Correlates of khat use during pregnancy: A cross-sectional study. Addict Behav. 2017;73:178–84.

14. Mekuriaw B, Belayneh Z, Yitayih Y. Magnitude of Khat use and associated factors among women attending antenatal care in Gedeo zone health centers, southern Ethiopia: a facility based cross sectional study. BMC Public Health. 2020;20(1):110.

15. Yadeta TA, Egata G, Seyoum B, Marami D. Khat chewing in pregnant women associated with prelabor rupture of membranes, evidence from eastern Ethiopia. Pan Afr Med J. 2020;36:1–.

16. Fetene MT, Teji K, Assefa N, Bayih WA, Tsehaye G, Hailemeskel HS. Magnitude and associated factors of substance use among pregnant women attending antenatal care in public hospitals of eastern Ethiopia. BMC Psychiatry. 2021;21(1):96.

17. Tesso FY, Woldesemayat L, Kebede DB. Magnitude of substance use and associated factors among pregnant women attending jimma town public health facilities, Jimma Zone, Oromia Regional State Southwest Ethiopia. Clinics Mother Child Health. 2017;14(275):2.

18. Tesfay K, Abera M, Wondafrash M, Tesfaye M. Effect of Khat Use During Pregnancy on the Birth Weight of Newborn in Jimma, Ethiopia. International Journal of Mental Health and Addiction. 2019;17(6):1432–41.

19. Kassim S, Jawad M, Croucher R, Akl EA. The Epidemiology of Tobacco Use among Khat Users: A Systematic Review. Biomed Res Int. 2015;2015:313692.

20. Alem A, Kebede D, Kullgren G. The prevalence and socio-demographic correlates of khat chewing in Butajira, Ethiopia. Acta Psychiatr Scand Suppl. 1999;397:84–91.

21. Alemu WG, Amare Zeleke T, Takele WW. Prevalence and associated factors of khat chewing among students in Ethiopia: a protocol for systematic review and meta-analysis. BMJ Open. 2018;8(11):e021157.

22. Demelash H, Motbainor A, Nigatu D, Gashaw K, Melese A. Risk factors for low birth weight in Bale zone hospitals, South-East Ethiopia : A case-control study. BMC Pregnancy and Childbirth. 2015;15(1).

23. Mekonnen AG, Hordofa AG, Kitila TT, Sav A. Modifiable risk factors of congenital malformations in bale zone hospitals, Southeast Ethiopia: an unmatched case-control study. BMC Pregnancy Childbirth. 2020;20(1):129.

24. Manzar MD, Salahuddin M, Sony P, Maru TT, Pandi-Perumal SR, Moscovitch A, et al. Sleep disturbances and memory impairment among pregnant women consuming khat: An under-recognized problem. Annals Of Thoracic Medicine. 2017;12(4):247–51.

25. Shay JW, Homma N, Zhou R. Abstracts from the 3rd International Genomic Medicine Conference (3rd IGMC 2015) : Jeddah, Kingdom of Saudi Arabia. 30 November - 3 December 2015. BMC Genomics. 2016;17 Suppl 6(Suppl 6):487.

26. Kedir H, Berhane Y, Worku A. Khat chewing and restrictive dietary behaviors are associated with anemia among pregnant women in high prevalence rural communities in eastern Ethiopia. Plos One. 2013;8(11):e78601–e.

27. Muhammed SAM, Muhammed, A.K. Al-Mansoob. The Impact of Chewing Khat during Pregnancy on Foetal Death History. International Journal of Novel Research in Healthcare and Nursing Novelty Journals. 2015;2(2):28–31.

28. Abdelaleem M. Khat Chewing During Pregnancy: an Insight on an Ancient Problem Impact of Chewing Khat on Maternal and Fetal Outcome among Yemeni Pregnant Women. Journal of Gynecology & Neonatal Biology. 2016;1:1–4.

29. Testa J. The Thomson Reuters journal selection process. Transnational Corporations Review. 2009;1(4):59–66.

30. Liberati A, Altman DG, Tetzlaff J, Mulrow C, Gøtzsche PC, Ioannidis JPA, et al. The PRISMA statement for reporting systematic reviews and meta-analyses of studies that evaluate health care interventions: explanation and elaboration. PLoS medicine. 2009;6(7):e1000100–e.

31. Peterson J, Welch V, Losos M, Tugwell P. The Newcastle-Ottawa scale (NOS) for assessing the quality of nonrandomised studies in meta-analyses. Ottawa: Ottawa Hospital Research Institute. 2011:1–12.

32. Stang A. Critical evaluation of the Newcastle-Ottawa scale for the assessment of the quality of nonrandomized studies in meta-analyses. Eur J Epidemiol. 2010;25(9):603–5.

33. Higgins JP, Thompson SG, Deeks JJ, Altman DG. Measuring inconsistency in meta-analyses. Bmj. 2003;327(7414):557–60.

34. Woldetsadik AM, Ayele AN, Roba AE, Haile GF, Mubashir K. Prevalence of common mental disorder and associated factors among pregnant women in South-East Ethiopia, 2017: a community based cross-sectional study. Reprod Health. 2019;16(1):173.

35. Mekuriaw B, Belayneh Z, Shemelise T, Hussen R. Alcohol use and associated factors among women attending antenatal care in Southern Ethiopia: a facility based cross sectional study. BMC Res Notes. 2019;12(1):690.

36. Alamneh AA, Endris BS, Gebreyesus SH. Caffeine, alcohol, khat, and tobacco use during pregnancy in Butajira, South Central Ethiopia. PLoS One. 2020;15(5):e0232712.

37. Tesfaye G, Demlew D, M GT, Habte F, Molla G, Kifle Y, et al. The prevalence and associated factors of alcohol use among pregnant women attending antenatal care at public hospitals Addis Ababa, Ethiopia, 2019. BMC Psychiatry. 2020;20(1):337.

38. Naegle MA. Brief Report: First World Health Organization Forum on Alcohol Drugs and Addictive Behaviors: Enhancing Public Health Actions Through Partnerships and Collaboration. Journal of Addictions Nursing. 2017;28(3):150–1.

39. Wondemagegn AT, Cheme MC, Kibret KT. Perceived Psychological, Economic, and Social Impact of Khat Chewing among Adolescents and Adults in Nekemte Town, East Welega Zone, West Ethiopia. BioMed Research International. 2017;2017:7427892.

40. Tessema ZT, Zeleke TA. Spatial Distribution and Factors Associated with Khat Chewing among Adult Males 15-59 Years in Ethiopia Using a Secondary Analysis of Ethiopian Demographic and Health Survey 2016: Spatial and Multilevel Analysis. Psychiatry J. 2020;2020:8369693.

41. Onah MN, Field S, van Heyningen T, Honikman S. Predictors of alcohol and other drug use among pregnant women in a peri-urban South African setting. International Journal of Mental Health Systems. 2016;10(1):38.

42. Adebowale OO, James BO. Psychoactive substance use and psychiatric morbidity among pregnant women attending an ante-natal clinic in Benin City, Nigeria. Niger Postgrad Med J. 2018;25(1):8–12.

43. O’Keeffe LM, Kearney PM, McCarthy FP, Khashan AS, Greene RA, North RA, et al. Prevalence and predictors of alcohol use during pregnancy: findings from international multicentre cohort studies. BMJ Open. 2015;5(7):e006323.

44. Yitayih Y, van Os J. Prevalence and determinants of chewing khat among women in Ethiopia: data from Ethiopian demographic and health survey 2016. BMC psychiatry. 2021;21(1):1–8.

45. Gebrie A, Alebel A, Zegeye A, Tesfaye B. Prevalence and predictors of khat chewing among Ethiopian university students: A systematic review and meta-analysis. PLOS ONE. 2018;13(4):e0195718.

46. Organization WH. mhGAP intervention guide for mental, neurological and substance use disorders in non-specialized health settings: mental health Gap Action Programme (mhGAP) : World Health Organization; 2016.

